# Genomic epidemiology reveals geographical clustering of multidrug-resistant *Escherichia coli* sequence type (ST)131 associated with bacteraemia in Wales, United Kingdom

**DOI:** 10.1101/2021.05.21.21257487

**Authors:** Rhys T. White, Matthew J. Bull, Clare R. Barker, Julie M. Arnott, Mandy Wootton, Lim S. Jones, Robin A. Howe, Mari Morgan, Melinda M. Ashcroft, Brian M. Forde, Thomas R. Connor, Scott A. Beatson

## Abstract

2.

Increasing resistance to third-generation cephalosporins (3GCs) threatens public health, as these antimicrobials are prescribed as empirical therapies for systemic infections caused by Gram-negative bacteria. Resistance to 3GCs in urinary tract infections (UTIs) and bacteraemia is associated with the globally disseminated, multidrug-resistant, uropathogenic *Escherichia coli* sequence type (ST)131. This study combines the epidemiology of *E*.*coli* blood culture surveillance with whole-genome sequencing (WGS) to investigate ST131 associated with bacteraemia in Wales between 2013 and 2014. This population-based prospective genomic analysis investigated temporal, geographic, and genomic risk factors. To identify spatial clusters and lineage diversity, we contextualised 142 genomes collected from twenty hospitals, against a global ST131 population (*n*=181). All three major ST131 clades are represented across Wales, with clade C/*H*30 predominant (*n*=102/142, 71.8%). Consistent with global findings, Welsh strains of clade C/*H*30 contain *β*-lactamase genes from the *bla*_CTX-M-1_ group (*n*=65/102, 63.7%), which confers resistance to 3GCs. In Wales, the majority of clade C/*H*30 strains belonged to sub-clade C2/*H*30Rx (*n*=88/151, 58.3%), whereas sub-clade C1/*H*30R strains were less common (*n*=14/67, 20.9%). A sub-lineage unique to Wales was identified within the C2/*H*30Rx sub-clade (named GB-WLS.C2/*H*30Rx) and is defined by six non-recombinogenic single-nucleotide polymorphisms (SNPs), including a missense variant in *febE* (ferric enterobactin transport protein) and *fryC* (fructose-like permease IIC component), and the loss of the capsular biosynthesis genes encoding the K5 antigen. Bayesian analysis predicted that GB-WLS.C2/*H*30Rx diverged from a common ancestor (CA) most closely related to a Canadian strain between 1998 and 1999. Further, our analysis suggests a descendent of GB-WLS.C2/*H*30Rx arrived through an introduction to North Wales circa 2002, spread and persists in the geographic region, causing a cluster of cases (CA emerged circa 2009) with a maximum pair-wise distance of 30 non-recombinogenic SNPs. This limited genomic diversity likely depicts local transmission within the community in North Wales. This investigation emphasises the value of genomic epidemiology, allowing detection of suspected transmission clusters and the spread of genetically similar/identical strains in local areas. These analyses will enable targeted and timely public health interventions.

**Impact statement:** Uropathogenic *Escherichia coli* (UPEC) is a leading cause of bacteraemia, resulting in substantial mortality and morbidity, with rates of *E. coli* bacteraemia (ECB) becoming a particular concern in Wales(1). Previous genomic and multilocus sequence typing (MLST) studies have identified that ECB cases are disproportionately caused by specific groups [sequence types (ST)] of related *E. coli*. Previous work reports ST131 as a globally disseminated lineage associated with bacteraemia and antimicrobial resistance (AMR). Despite widespread study of ECB, the temporal and geographic patterns of key ECB clones remain an important area of study. Moreover, by gaining a detailed understanding of the population structure of key ECB clones, it should be possible to develop and improve public health measures to reduce the risk of ECB and act to combat the rise of AMR. Using whole-genome sequencing, we describe the temporal and spatial relationship of a collection of *E. coli* ST131 bacteraemia cases sampled across Wales. High-resolution analyses of genetic variants identified a local (North Wales) cluster of strains within the highly antimicrobial-resistant sub-clade C2/*H*30Rx, which are characterised by resistance to nitrofurantoin and the loss of the K5 capsule. Notably, AMR stewardship guidelines in Wales recently changed to include nitrofurantoin as a first-line treatment for uncomplicated UTIs. This local cluster likely represents environmentally-mediated community transmission, environmentally mediated, from the strain’s common ancestor that existed circa 2009, highlighting the need for national genomic surveillance, close to real-time, to track and understand the evolution of AMR in communities.

**Data summary:** The study sequences are available in the National Center for Biotechnology Information (NCBI) under BioProject accession number PRJNA729115. Raw Illumina sequence read data have been deposited to the NCBI sequence read archive [SRA (https://www.ncbi.nlm.nih.gov/sra)] under the accession numbers SRR14519411 to SRR14519567. A complete list of SRA accession numbers is available in Table S1 (available in the online version of this article). The high-quality draft assemblies have been deposited to GenBank under the accession numbers JAHBGJ000000000 to JAHBMG000000000, and JAHBRR000000000 to JAHBRT000000000. The programs used to analyse raw sequence reads for polymorphism discovery and whole-genome sequencing based phylogenetic reconstruction are available as described in the materials and methods. The authors confirm all supporting data, code, and protocols have been provided within the article or through supplementary data files.

## 5. Introduction

Uropathogenic *Escherichia coli* (UPEC), bacteria causing infection rather than commensal bacteria, are the leading cause of urinary and systemic infections. UPEC present an increasing burden to public health due to increasing antimicrobial resistance (AMR). Increasing rates of AMR can lead to treatment failures and progression to systemic bacteraemia infections. Additionally, the emergence and dissemination of UPEC strains encoding AMR are causing economic damage to countries and healthcare systems(2). Incidences of *E. coli* associated bacteraemia are increasing globally. In Wales, the 5-year rolling average age-standardised mortality for deaths involving *E. coli* bacteraemia (ECB) almost doubled from 4.0 [95% confidence interval (CI): 2.3 to 6.4] per 1 million population in 2002-06, to 7.7 (95% CI: 5.4 to 10.6) in 2006-10 (Figure S1)(1). Previous studies have shown that most urinary tract infections (UTIs) are caused by a limited number of key UPEC clonal lineages including sequence types (ST)131, ST69, ST73, ST95 and ST12(3-5). UPEC are predominantly found within the *E. coli* phylogenetic groups B1, B2, or D.

*E. coli* ST131 is a high-risk pandemic clone that is frequently associated with bacteraemia(6) and UTIs, and is a major circulating lineage in the United Kingdom (UK)(7). The successful transmission of ST131 globally is attributed to: (i) resistance to many treatments by the carriage of genes encoding resistance to multiple antimicrobial agents(8-10); (ii) the ability to cause disease that other opportunistic or commensal strains do not possess through pathogenicity, fitness, and metabolic factors(11-13); (iii) the ability to survive in human serum(14) due to capsule production(15); and (iv) transmission in various environments including healthcare- and community-acquired transmission(16). ST131 can colonise and persist in hosts for extended periods causing recurrent UTIs, typically within 1-year of the initial infection(17). ST131 cases frequently harbour resistance to many broad-spectrum therapies such as third-generation cephalosporins (3GCs)(18, 19) and fluoroquinolones(20). In ST131, the carriage of extended-spectrum *β*-lactamases (ESBLs) facilitates the principal resistance mechanism to 3GCs.

In 2018, the UK National Institute for Clinical Excellence (NICE) issued guidelines concerning acute pyelonephritis(21). This recommended urine culture susceptibility testing and promoted the use of several *β*-lactams, trimethoprim, ciprofloxacin (fluoroquinolone), or amoxicillin and clavulanic acid as first-line antibiotics. This could be contributing to increasing rates of ESBL-producing *E. coli* across the UK(22). Between 2017-2018, data from England and Wales showed that at least 14.1% (*n*=4,950/35,050)(23) and 13.3% (*n*=354/2,663)(24) of *E. coli* bloodstream isolates presented resistance to 3GCs, respectively. In England, this translates to approximately 5,000 annual cases, often due to ST131(4, 7). Resistance to *β*-lactams like 3GCs can lead to increased usage of last-line therapies like carbapenems, with carbapenem resistance in UPEC also associated with ST131(25, 26). In 2017, rates of resistance to fluoroquinolones in ECB cases across Wales were as at least 20.3% (*n*=540/2,663)(24). NICE also promotes the use of nitrofurantoin or trimethoprim (first-line), and pivmecillinam or fosfomycin (second-line) antibiotics against lower UTIs(27). However, trimethoprim is no longer recommended in Wales for the treatment of UTIs in the 65 and over age group(24). The increase in antimicrobial-resistant infections is problematic on several levels. For example, patients are more likely to receive inappropriate empirical therapy involving an agent to which the pathogen is resistant. The circulation of strains with extensive levels of resistance to key antimicrobials, such as 3GCs, increases the likelihood of UTI treatment failures, prolonging the length of infection, potentially allowing the bacteria to flourish by removing commensal bacteria which compete for bacterial growth, and increases severe outcomes such as the risk of a patient developing bacteraemia, resulting in increased morbidity and mortality.

Genomic epidemiology, the use of whole-genome sequencing (WGS) in epidemiological investigations, is increasing worldwide in public health responses. With increasing rates of antimicrobial-resistant ECB, it is vital to understand the genetic relatedness of circulating strains on a local, national, and global scale. This work investigated the evolution of ST131 strains from patients in Wales with bacteraemia identified over a 12-month period. Genomic sequence data enabled the characterisation of circulating ST131 in Wales, showing multiple introductions of this global clone with localised or national transmission. Our analyses also reveal multiple bacteraemia cases caused by a unique geographically-restricted, monophyletic subgroup of ST131 within North Wales characterised by ESBL-production.

## 6. Methods

### 6.1 Welsh *E. coli* isolate collection and genome sequencing

Public Health Wales (PHW) laboratories were asked to submit all *E. coli* blood isolates from blood samples collected between April 2013 and March 2014, to the national Specialist Antimicrobial Chemotherapy Unit (SACU) at University Hospital Wales. The isolate dataset was linked to routine microbiological surveillance data by PHW to obtain isolate AMR profiles. Novel AMR profiles and profiles with phylo-geography were characterised by polymerase chain reaction. Selected samples were sequenced based on their determined phylogenetic groups. Isolates were transported to Cardiff University, cultured overnight in liquid culture, and extracted using a Promega (Wisconsin, USA) Maxwell instrument. Samples were sequenced as paired-end reads on either the NextSeq 500 or HiSeq 2500 platform (Illumina Inc, San Diego, CA, USA) at the Oxford Genomics Centre (https://www.well.ox.ac.uk/ogc/) or MicrobesNG (https://microbesng.com/). DNA libraries were prepared using a mixture of the Nextera XT® Library Preparation Kit and the NEBNext® Ultra™ Library Preparation Kit (Illumina Inc, San Diego, CA, USA), following the manufacturer’s instructions in both cases.

This genomic surveillance of ECB in Wales collected 157 non-duplicate clinical *E. coli* ST131 strains as part of routine microbiological surveillance data from hospitals across six administrative units known as health boards (Figure S2). Patient anonymity was maintained by pseudonymised data that went outside PHW. Epidemiological information, including isolate names and available metadata are summarised in Supplementary Tables S1 and S2. The WGS of the 157 *E. coli* isolates generated a median of 0.89 million paired-end reads per sample [interquartile range (IQR): 0.43 to 1.09 million; range: 0.14 to 3.37 million] (Table S2). Sequence read data for all Welsh isolates were submitted to the National Center for Biotechnology Information Sequence Read Archive under BioProject accession number PRJNA729115. The methods used for quality control for this dataset are available in the Supplementary Methods. Briefly, we identified and excluded the sequence data for 15 isolates from further analysis based on the sequencing coverage below 20-fold (Table S3).

### 6.2 Draft genome assembly

Quality-trimmed paired-end reads for the remaining 142 Welsh strains were *de novo* assembled using MGAP (https://github.com/dsarov/MGAP---Microbial-Genome-Assembler-Pipeline), which implements: Velvet v1.2.10(28); VelvetOptimiser (https://github.com/tseemann/VelvetOptimiser); GapFiller v1.10(29); ABACAS v1.3.1(30) [scaffolds against the chromosome of *E. coli* ST131 strain EC958 (GenBank: HG941718)]; IMAGE v2.4(31); SSPACE v2.0(32); Pilon v1.22(33); and MIRA v4(34). Contigs from the draft assemblies were ordered against the complete chromosome of EC958 using Mauve version snapshot_2015-02-25(35). QUAST v4.5(36) assessed the assembly statistics generated from MGAP by comparing each isolate to EC958 (Table S4).

### 6.3 Complementary datasets

To facilitate the geographic analysis of the 142 Welsh ST131 isolates within the global context, available isolate datasets were downloaded including: (i) sequence read data from the NCBI sequence read archive (SRA); (ii) draft assemblies; and (iii) associated metadata from Ben Zakour *et al*.(10) (*n*=189) and Kidsley *et al*.(37) (*n*=19). Notably, six draft assemblies from the Ben Zakour *et al*. study were replaced with the complete chromosomes: CD306 (GenBank: CP013831); JJ1886 (GenBank: CP006784); JJ1887 (GenBank: CP014316); JJ2434 (GenBank: CP013835); S65EC (GenBank: CP036245); and ZH193 (GenBank: CP014497) (Table S5). The methods used for *in silico* gene typing and generation of an assembly based ST131 phylogeny (initial context) for this global dataset are available in the Supplementary Methods.

### 6.4 Compiling a high-quality ST131 clade C/*H*30 global dataset

For context, the Welsh clade C/*H*30 strains (*n*=102) were inputted against a global collection of clade C/*H*30 ST131 strains (*n=*117) from three published studies(9, 10, 38) as featured in Kidsley *et al*.(37). Several complete genomes were integrated by simulating error free reads using ART (version ART-MountRainier-2016-06-05)(39) to 60x coverage with an insert size of 340 ± 40 bp. These included known clade C/*H*30 ST131 genomes: 2/0 (GenBank: CP023853); 4/0 (GenBank: CP023849); 4/4 (GenBank: CP023826); 4/1-1 (GenBank: CP023844); MNCRE44 (GenBank: CP010876); U12A (GenBank: CP035476); U13A (GenBank: CP035477); U14A (GenBank: CP035516); U15A (GenBank: CP035720); and uk_P46212 (GenBank: CP013658). For this investigation, clade C/*H*30 strains JJ2183 (SRA: SRS456889), MVAST0036 (SRA: SRS456851), MVAST046 (SRA: SRS456881), and MVAST077 (SRA: SRS456882) were removed from the dataset based on the average sequence coverage depth below 20-fold.

### 6.5 Identifying genetic variants

High-resolution analyses of genetic variants was performed using the Burrows– Wheeler Aligner v0.7.15(40); SAMtools v1.2(41); Picard v2.7.1 (https://github.com/broadinstitute/picard); the Genome Analysis Tool Kit v3.2-2 (GATK)(42, 43); BEDTools v2.18.2(44); and SNPEff v4.1(45) as implemented in SPANDx v3.2(46). In brief, the trimmed reads were mapped to the complete chromosome of EC958; which was isolated in March 2005 in the United Kingdom from a community-onset urine infection in an 8-year-old girl(11). When analysing the Illumina reads, 20 clade C/*H*30 genomes from Price *et al*.(38) were excluded from this example dataset due to erroneous Phred quality encoding (Table S6). Our final dataset consisted of 226 genomes representing previously published datasets (*n*=124, including 16 complete genomes) and our Welsh collection (*n*=102) (Table S7).

### 6.6 High-resolution phylogeny of the clade C/*H*30 ST131 sub-lineage

The 226 strains [EC958 reference (*n*=1) and dataset (*n*=225)] were assessed for the presence of strain mixtures (see Supplementary Methods). Briefly, this approach readily flagged seven strains as a probable mixture based on the high number of ambiguous single-nucleotide polymorphisms (SNPs) (from a total of 7,592 SNPs) compared with the remaining 219 genomes [Median 51 (0.7%); IQR 28 to 102 (0.4 to 1.3%); range 2 to 171 (0.0 to 2.3 %)]. The quality-trimmed paired-end Illumina reads from the remaining 218 high-quality clade C/*H*30 isolates were mapped onto the chromosome of EC958 using SPANDx with default parameters to generate annotated SNP/INDEL matrices. To account for recombination, regions of high-density clustered SNPs (≥3 SNPs found within a 100 bp window) were removed. Sites were excluded if a SNP was called in regions with less than half or greater than 3-fold the average genome coverage on a genome-by-genome basis. SPANDx generated an alignment of 4,354 non-recombinant, orthologous, biallelic core-genome SNPs from the 219 strains. Lastly, a maximum likelihood (ML) phylogenetic tree from the non-recombinant SNP alignment was generated using RAxML v8.2.10(47) (GTR-GAMMA correction) thorough optimisation of 20 distinct, randomized maximum parsimony trees, before adding 1,000 bootstrap replicates. The resulting phylogenetic tree was visualised using FigTree v1.4.4 (http://tree.bio.ed.ac.uk/software/figtree/) and EvolView v2(48, 49).

### 6.7 Root-to-tip regression analysis

A regression analysis was used to estimate the temporal signal in the clade C/*H*30 ST131 sub-lineage between the root-to-tip genetic distance using TempEst v1.5.15(50). The ML phylogenetic tree that was reconstructed from the alignment of 4,354 non-recombinant, orthologous, biallelic core-genome SNPs (as described above) was used as the input into TempEST. Four problematic sequences that did not match the evolutionary trajectory of the remaining strains within the lineage were identified and were removed from the temporal analysis [BA1581 (SRA: SRR14519500), JJ2643 (SRA: SRS456891), S77EC (SRA: ERR161304), and U004 (SRA: SRS456902)]. SPANDx was rerun using the same parameters as described above, only with the ‘-s’ flag set to none to move straight to the comparative genomics and error correction section of the pipeline. The new alignment for Bayesian phylogenetic inference consisted of 4,150 non-recombinant, orthologous, biallelic core-genome SNPs from 215 clade C/*H*30 ST131 strains. The ML rebuilt using the methods above. Again, the tree was rooted by CD306.

### 6.8 Bayesian temporal analysis

To further the temporal analysis, a time calibrated phylogenetic tree was generated with BEAST2 v2.6.1(51). The alignment of 4,150 SNPs was run through jModelTest v2.1.10(52, 53), which identified the GTR nucleotide substitution model as the best-fit evolutionary model. To test if the strict clock or uncorrelated relaxed clock best-fit our dataset, initial models were created using tip dates, a GTR substitution model, and a coalescent prior with a constant population. Both models were tested with the Nested sampling Bayesian computation algorithm v1.1.0 within the BEAST2 package with a particle count of 1, sub chain length of 5,000, and Epsilon of 1.0×10^−6^. This analysis provides evidence in favour of the uncorrelated relaxed clock model. Various population models were compared to ensure selection of the best-fit model. These included the Bayesian skyline, coalescent constant, and exponential growth population size change models. The Gamma Site Model Category Count was set to four and the GTR substitution model rates determined from jModelTest were included (i.e., rate AC = 0.94, AG = 3.16, AT = 1.10, CG = 0.14, CT = 3.12, and GT =1.00). Notably, the initial clock rate was set to 7.61×10^−4^ (as estimated from the root-to-tip regression analysis in TempEST) with a uniform distribution and an upper bound of 0.1. All other priors were left as default. A total of three independent Markov chain Monte Carlo (MCMC) generations for each analysis were conducted for 100 million generations. Trees were sampled every 1,000 generations which resulted in triplicate samples of 100,000 trees for each model test. All BEAST runs were imported into Tracer v1.7.1 (http://github.com/beast-dev/tracer/) to assess statistics. LogCombiner v2.5.0 (BEAST 2 package) then combined the replicated analyses for each model with a 10% burn-in to assess convergence/appropriate sampled run. Finally, TreeAnnotator v2.4.5 (BEAST 2 package) removed the 10% burn-in and generated maximum clade credibility (MCC) trees for each run (established from 243 million trees), reporting median values with a posterior probability limit set at 0.5. FigTree was used to visualise the annotated MCC trees. We determined the best-fitting tree model as the uncorrelated relaxed exponential clock model with the Bayesian skyline population size change model based on the mean tree likelihood scores (Table S8).

## 7. Results

### 7.1 *E. coli* ST131 strains from Wales

Previous genomic and multilocus sequence typing (MLST) investigations identified that ECB cases were disproportionately caused by *E. coli* ST131 (*n*=187/720, 26.0%)(54). This study involved *E. coli* ST131 isolates cultivated from blood specimens collected from twenty hospitals across six health boards within Wales (Figure S2). Of the 157 isolates collected over the study (between 2013 and 2014), 142 passed quality-control on the sequence data [females *n*=70/142 (49.3%), males *n*=69/142 (48.6%), no sex recorded *n*=3/142 (2.1%)]. Patients were typically older, with a median age of 80 years (IQR: 70 to 87 years; range: 19 to 105 years), which reflects the known patient profile of ECB cases(1).

### 7.2 Major ST131 clades are represented amongst ST131 circulating Wales

The 142 draft genomes had a median total length of 5.20 Mb (IQR: 5.10 to 5.27; range: 4.75 to 5.48 Mb), a median GC content of 50.7% (IQR: 50.7 to 50.8%; range: 50.5 to 51.3%), and a median N50 statistic of 199.59 kb (IQR: 102.39 to 245.60 kb; range: 6.42 to 499.50 kb). All 142 genomes are ST131, except for BA1243 and BA1279 (same Clonal Complex) that differ in the fumarate hydratase class II (*fumC*) and malate dehydrogenase (*mdh*) genes respectively (Figure S3). The Welsh ST131 draft genomes have a high prevalence of chromosomal mutations conferring high level resistance (MICs >32mg/L) to fluoroquinolones, where most (*n*=102/142, 71.8%) contain double variants in *gyrA* (D87N and S83L) and *parC* (E84V & S80I). An additional ten genomes contain a single variant in *gyrA* (S83L) which usually confers low level resistance (MICs 0.5mg/L). The *bla*_CTX-M-15_ gene (CTX-M-1 group) is the most common (*n*=65/142, 45.8%) AMR gene encoding ESBLs. The *bla*_OXA-1_ gene is also common (*n*=62/142, 43.7%) amongst Welsh strains, which encodes resistance to amoxicillin/clavulanic acid, and piperacillin/tazobactam (antibiotic/*β*-lactamase inhibitor when in combination with ESBL genes). Notably, 36.6% (*n*=52/142) of the strains carry both *bla*_CTX-M-15_ and *bla*_OXA-1_. To capture a snapshot of the genomic diversity and population structure amongst ST131 strains circulating in Wales, the draft assemblies of the 142 genomes were contextualised with a global collection of ST131 cases sequenced elsewhere (*n*=208) (Figure S4). The 13,758 non-recombinant core-genome SNP alignment represents a core-genome alignment of 2,575,140 bp relative to the 5,109,767 bp reference chromosome EC958. All three well-supported major ST131 clades (A, B, and C) are represented across Wales. While most isolates are located within clade C/*H*30 (*n*=103/142, 72.5%), representatives from clade A (*n*=22/142, 15.5%) and clade B (*n*=17/142, 12.0%) are present at similar frequencies.

### 7.3 The majority of clade C/*H*30 ST131 isolates circulating Wales are ESBL-producing, conferring resistance to 3GCs

To infer the phylogenetic relatedness of isolates and determine AMR gene carriage, we created a core-genome SNP alignment of clade C/*H*30 strains only using SPANDx. This alignment of global clade C/*H*30 strains (*n*=219) comprises 4,354 non-recombinant orthologous biallelic SNPs representing a ∼4,005,300 bp core-genome (regions estimated to the nearest 100 bp with ≥95% coverage across all genomes) relative to the 5,109,767 bp chromosome of EC958. Almost half (*n*=102/219, 46.6%) of our global clade C/*H*30 lineage comprised isolates collected from bacteraemia cases across Wales (Figure 1; see branch lengths expressed as SNPs in Figure S5). In our combined dataset, the majority of clade C/*H*30 ST131 strains belonged to sub-clade C2/*H*30Rx (*n*=151/219, 68.9%), with sub-clade C1/*H*30R (*n*=67/219, 30.6%) less common. Sub-clade C2/*H*30Rx mostly comprised Welsh strains (*n*=88/151, 58.3%), whereas in sub-clade C1/*H*30R, the Welsh strains comprise only 20.9% of the sub-lineage (*n*=14/67). The majority (*n*=68/102, 66.7%) of these clade C/*H*30 isolates demonstrate an ESBL-producing genotype. In terms of acquired resistance to *β*-lactams, CTX-M-type metallo-*β*-lactamase genes were dominant, with the most prevalent being the *bla*_CTX-M-1_ group (*n*=65/102, 63.7%). The second most prevalent *β*-lactamase (*n*=61/102, 59.8%) *bla*_OXA-1_, which encodes resistance to amoxicillin/clavulanic acid, and piperacillin/tazobactam (antibiotic/*β*-lactamase inhibitor).

**Figure 1.**
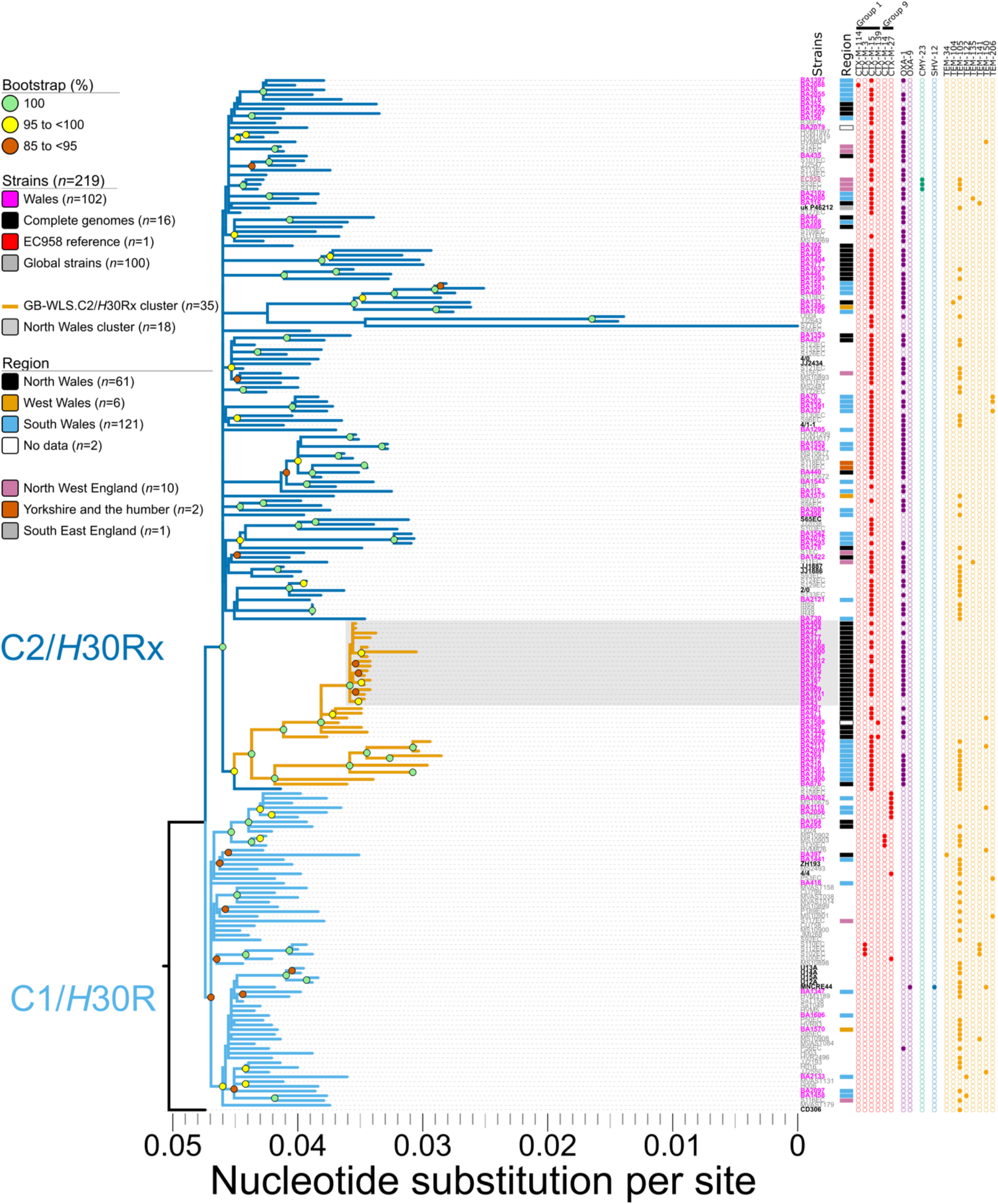
Maximum likelihood phylogeny of clade C/*H*30 *Escherichia coli* sequence type (ST)131 isolates plotted against *β*-lactam resistance complement. Phylogeny inferred from 4,354 non-recombinant orthologous biallelic core-genome single-nucleotide polymorphisms (SNPs) from 219 strains. Moderate recombination SNP density filtering in SPANDx (excluded regions with ≥3 SNPs in a 100 bp window). SNPs are derived from read mapping to the reference chromosome EC958 (GenBank HG941718). Phylogenetic trees are rooted according to the CD306 (GenBank: CP013831) outgroup. Branch lengths represent nucleotide substitutions per site as indicated by the scale bar. Bootstrapping using 1,000 replicates demonstrates the robustness of the branches.

### 7.4 The GB-WLS.C2/*H*30Rx Welsh cluster shares a common ancestor of North American origin

Among the C2/*H*30Rx population (Figure 1), there is a cluster of 35 isolates from Wales that are separated by a maximum pair-wise distance of 123 non-recombinogenic SNPs between strains BA264 (collected in South Wales in 2013) and BA2000 (collected in North Wales in 2014). Strains within this C2/*H*30Rx Welsh cluster (designated GB-WLS.C2/*H*30Rx) are closely related with a median pair-wise distance of 48 (IQR: 21 to 92) non-recombinogenic SNPs. The GB-WLS.C2/*H*30Rx sub-lineage has descended from a common ancestor (CA) shared with the clinical O25b:H4:K5 strain S125EC (SRA: ERS126605), which was cultivated in 2002 from a patient with a surgical wound in Canada(9), pointing towards the global dispersion of ST131. Isolates within GB-WLS.C2/*H*30Rx are distinguishable by six unique core-genome SNPs relative to EC958, two of which are in genes associated with fitness or virulence (*cusB*; cation efflux system mediating resistance to copper and silver and *fepE*; ferric enterobactin siderophore transport protein) (Table 1). Outside of the core-genome, strains within GB-WLS.C2/*H*30Rx (except for BA876) have lost region II of the K5 capsule loci, likely because of recombination, resulting in a loss of the K5 capsule production (Figure 2). In *E. coli*, group 2 capsular polysaccharides typically share conserved regions in the capsule loci (regions I and III). These conserved regions encode the transmembrane complex involved in the export and assembly of the capsular polysaccharides(15, 55, 56). Region II is, however, serotype-specific and encodes for enzymes responsible for synthesizing the capsular polysaccharide. Strains within GB-WLS.C2/*H*30Rx have region II of the capsule locus replaced with two genes, the *catB* chloramphenicol-related O-acetyltransferase (xenobiotic acyltransferase [XAT]), conferring resistance to chloramphenicol and a HAD-IA family hydrolase, which has been resolved with the completion of the clinical strain O25:H4 collected in Saudi Arabia in 2014 (GenBank: CP015085). In the C2/*H*30Rx strain O25:H4, the low average guanine-cytosine content (42.18%) of the 15.2 kb capsule locus suggests a foreign origin.

**Table 1.** Specific variants between *Escherichia coli* strain EC958 and GB-WLS.C2/*H*30Rx.

| Position in EC958 | EC958 | Cluster | Change<sup>b</sup> | Impact | Codon | Gene | Product |
| --- | --- | --- | --- | --- | --- | --- | --- |
| 601,912 | **C** | T | Syn | LOW | 383 | *cusB* | Cation efflux system protein |
| 632,160 | **C** | T | Q=>stop | HIGH | 130 | *fepE* | Ferric enterobactin transport |
| 1,672,896 | **T** | C | Syn | LOW | 130 | *dosP* | Oxygen sensor protein |
| 2,698,528 | **G** | A | T=>I | MODERATE | 232 | *fryC* | Fructose-like permease IIC component |
| 2,902,622 | **T** | C |  |  |  |  |  |
| 2,964,604 | **A** | G | Syn | LOW | 165 | *pncC* | Nicotinamide-nucleotide amidohydrolase |

Emboldened and italicised nucleotides are specific to sub-clade C2/H30Rx EC958 and isolates within the Welsh cluster, respectively
<sup>b</sup> Consequence of SNP relative to EC958 (C2/H30Rx). Synonymous change (Syn); non-synonymous changes to protein-coding genes are shown by single letter amino acid abbreviation (EC958 sequence on left, SNP impact on right); blank lines indicate variant in intergenic region

**Figure 2.**
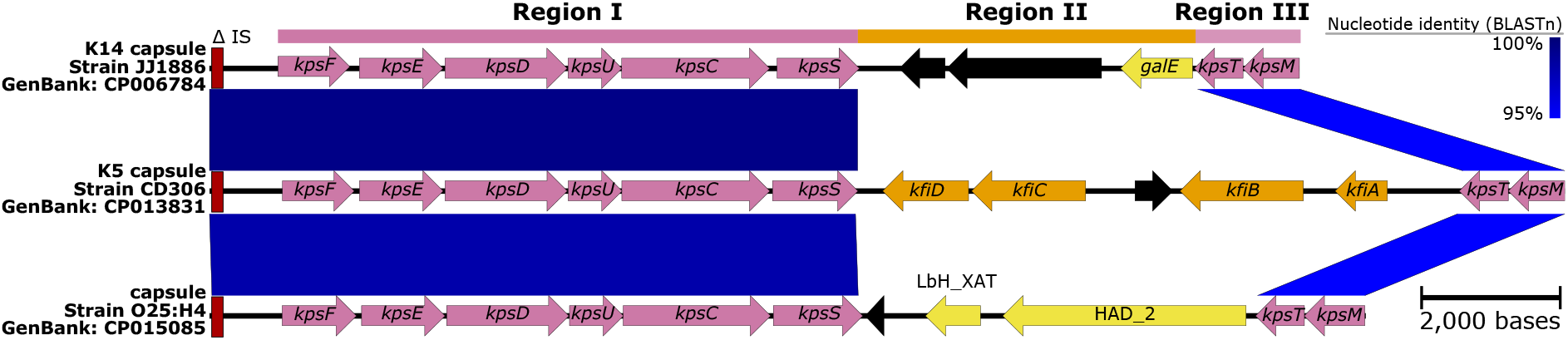
Major structural features and nucleotide pair-wise comparisons of the group 2 capsules in *Escherichia coli*. Nucleotide comparisons between the sub-clade C1/*H*30R genome JJ1886 and clade C/*H*30 outgroup genome CD306, highlighting differences between the K14 and K5 capsular region. Blue shading indicates nucleotide identity between sequences according to BLASTn (95 to 100%). Key genomic regions are indicated: IS: red (185bp fragment from IS110 family), conserved capsular regions I and III: pink, differing capsular regions (region II): orange and yellow, other CDSs: black. Image created using Easyfig(57).

Among GB-WLS.C2/*H*30Rx, there is a sub-cluster of 18 isolates from North Wales that are separated by a maximum pair-wise distance of 30 (median: 11, IQR: 8 to 13) non-recombinogenic SNPs, highlighting a small, local ST131 cluster. Isolates within this GB-WLS.C2/*H*30Rx sub-cluster from North Wales are distinguishable by 10 unique SNPs and a single 1-bp deletion relative to EC958 (Table 2). Additionally, a single strain BA909 (collected in 2014), contained a SNP putatively conferring resistance to rifampicin in *rpoB* (Q513L)(58). Due to anonymisation of epidemiological data, it is unclear whether isolates BA434 and BA1512 (separated by 9 SNPs, collected 257 days apart) and BA810 and BA909 (separated by 12 SNPs, collected 90 days apart) were sampled from the same patient. This analysis however, identified two samples, BA43 and BA910 separated by a single SNP, from two different patients (both male, aged 87 and 77-years) from two different hospitals, 239 days apart. This sequence similarity may suggest that these cases are linked by transmission or were colonised/infected from the same source. Additionally, our data suggests transmission within a single hospital in this same North Wales sub-cluster For example, isolates BA408 and BA434 from two individual patients collected two days apart from within the same hospital are separated by only two SNPs, suggestive of an epidemiological link.

**Table 2.** Variants separating *Escherichia coli* strain EC958 and the North Wales sub-cluster.

| Position in EC958 | EC958 | Cluster | Change<sup>b</sup> | Impact | Codon | Gene | Product |
| --- | --- | --- | --- | --- | --- | --- | --- |
| 900,998 | **G** | A | W=>stop | HIGH | 159 | *nfsA* | Oxygen-insensitive NADPH nitroreductase |
| 1,052,210 | **G** | T | G=>C | MODERATE | 403 | *ycaQ* | Uncharacterized protein |
| 1,113,811 | **C** | T | Syn | LOW | 74 | *appB* | Cytochrome bd-II ubiquinol oxidase subunit 2 |
| 1,842,716 | **T** | C | S=>P | MODERATE | 2 | *ydiS* | Probable electron transfer flavoprotein-quinone oxidoreductase |
| 2,372,060 | **G** | A | Syn | LOW | 413 | *gatZ* | D-tagatose-1,6-bisphosphate aldolase subunit |
| 2,405,414 | **CG** | C | Deletion | MODIFIER |  |  |  |
| 2,468,793 | **G** | A | Syn | LOW | 92 | *yeiR* | Zinc-binding GTPase |
| 2,485,972 | **A** | G | Syn | LOW | 302 | *yejM* | Inner membrane protein |
| 3,196,673 | **G** | A | Syn | LOW | 258 | *xanQ* | Xanthine permease |
| 4,706,103 | **C** | G | W=>S | MODERATE | 31 | *mdtN* | Multidrug resistance protein |
| 4,734,158 | **A** | G | S=>P | MODERATE | 205 | *basS* | Sensor protein |

Emboldened and italicised nucleotides are specific to sub-clade C2/H30Rx EC958 and isolates within the North Wales sub-cluster, respectively
<sup>b</sup> Consequence of SNP relative to EC958 (C2/H30Rx). Synonymous change (Syn); non-synonymous changes to protein-coding genes are shown by single letter amino acid abbreviation (EC958 sequence on left, SNP impact on right); blank lines indicate variant in intergenic region

### 7.5 The root-to-tip distance is consistent with the BEAST2 temporal signal and phylogenetic reconstruction

The divergence time and evolutionary distance for the 215 clade C/*H*30 ST131 genomes (four strains were removed as they did not match the evolutionary trajectory of the remaining strains) showed a linear relationship (correlation coefficient=0.59), with the regression analysis in TempEST indicating that the genomes accumulate mutations at a rate of 7.61×10^−4^ substitutions per site per year (*R*^*2*^=0.35) (Figure S6). The time to the most recent common ancestor (MRCA) is estimated at the end of 1993 (95% confidence interval: 1989 to 1996). Likewise, BEAST2 pinpoints the time to MRCA to 1994 [95% highest posterior density (HPD): 1986 to 1998] (Figure 3) (based on median node height) and estimates a mutation rate of 6.87×10^−4^ substitutions per site per year (95% HPD: 5.10×10^−4^ to 8.60×10^−4^). This translates to 2.85 fixated SNPs per year per genome (95% HPD: 2.12 to 3.57), which means that approximately six SNPs can be expected to differ between two isolates sharing a MRCA one year back in time. A previous study highlighted the within-host diversity of ST131 residing in the intestinal tract of a single patient(17); where strains U13A and U14A were collected nine months apart. Here, these strains are depicted to be separated by seven SNPs. This analysis indicates that the CA to U13A and U14A existed approximately two (95% HPD: one to five) years prior to the initial collection of U13A in 2013, which gives credibility to our phylogenetic reconstruction. To correct for ascertainment bias, our dataset describes one SNP for every 963.9 bases across the ∼4 Mb core-genome. This translates to a genome-wide mutation rate of 7.15×10^−7^ mutations/year/site relative to genome size, which is consistent with previous large-scale temporal analyses of *E. coli* [4.39×10^−7^(10) and 4.14×10^−7^(59)] and *Shigella* [6.0×10^−7^ (60)].

**Figure 3.**
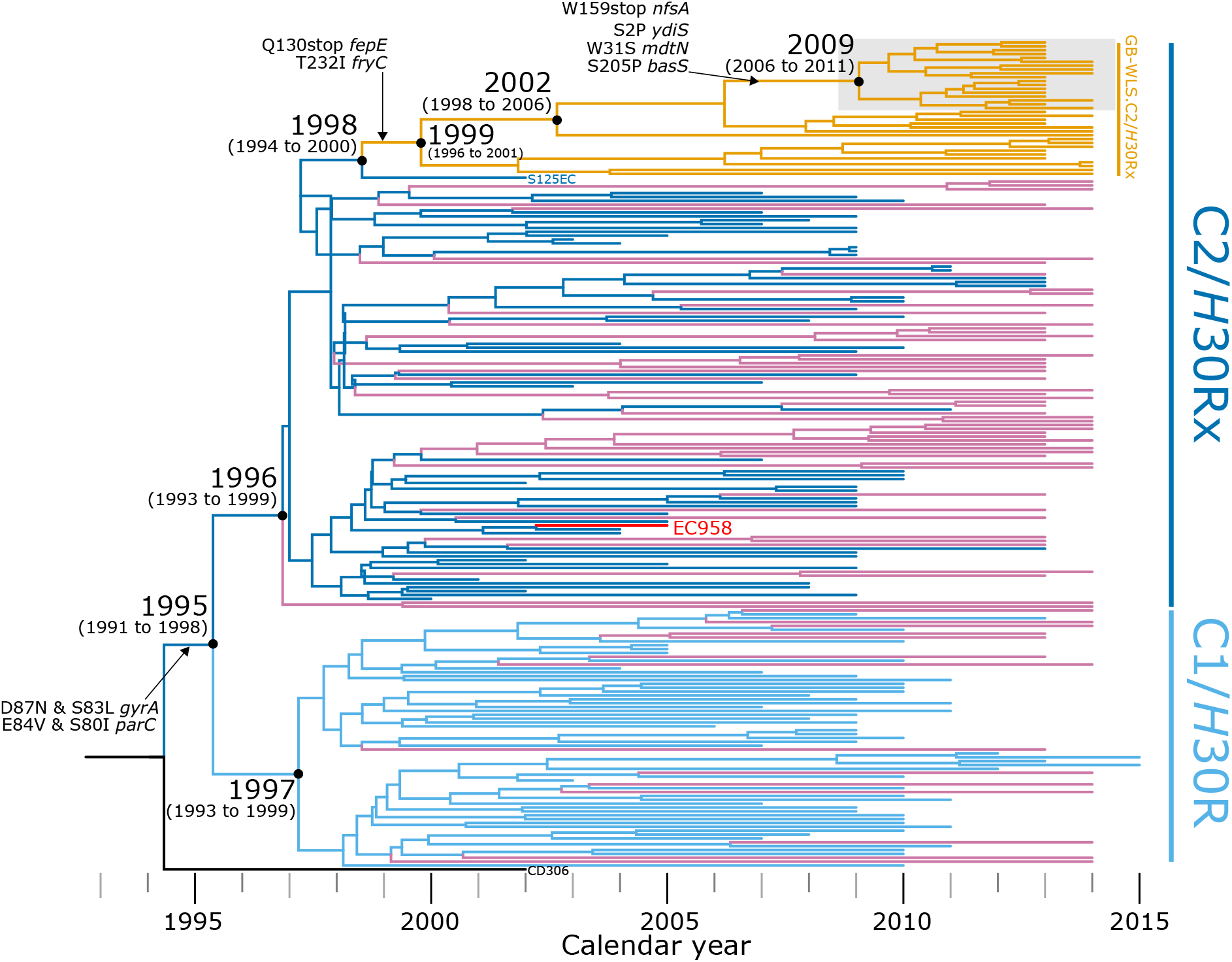
Evolutionary reconstruction of clade C/*H*30 *Escherichia coli* sequence type (ST)131. A time-calibrated maximum clade credibility tree inferred from 4,150 non-recombinant orthologous biallelic core-genome single-nucleotide polymorphisms (SNPs). Moderate recombination SNP density filtering in SPANDx (excluded regions with ≥3 SNPs in a 100 bp window). SNPs are derived from read mapping to the reference chromosome EC958 (GenBank: HG941718). X-axis represents the emergence time estimates. Isolates from Wales are shown with reddish purple or orange branches.

## 8. Discussion

Recent investigations across England have shown the value of identifying clonal lineages using high-resolution analyses obtainable through WGS for surveillance efforts(4, 7). For example, ECB in England is shown to represent a spill-over from strains circulating in the wider human population(4). This genomic epidemiology study provides a snapshot of the population structure of *E. coli* ST131 associated with bacteraemia in Wales. A final total of 142 *E. coli* ST131 strains collected across Wales underwent WGS and were contextualised for their genetic relationship with global ST131 strains through available datasets. To the knowledge of the study authors, this represents the first characterisation of the epidemiological and spatiotemporal nature of ST131 circulating in Wales.

This study used genomic epidemiology to identify clusters of strains, possibly suggesting patients were colonised/infected from the same source, and demonstrates the benefits of incorporating WGS, stringent quality control and epidemiological data for public health surveillance and investigations. Initial phylogenetic relationships were first inferred from SNPs to depict an overall tree topology that represent core-genome alignments of draft genome assemblies. These initial analyses found that, while all three major clades well-supported by the literature are represented in ST131 strains circulating in Wales, the predominant lineage is defined by a high prevalence of chromosomal mutations conferring resistance to fluoroquinolones and the presence of AMR genes encoding ESBLs (clade C/*H*30, particularly sub-clade C2/*H*30Rx). Subsequent analyses were undertaken to compile a much higher resolution of inter-strain relationships by exploiting read mapping of short-read sequence data to a chromosome previously sequenced. These analyses offer the opportunity for the re-evaluation of the ST131 clade C/*H*30 evolutionary trajectory previously characterised(4, 10, 59, 61) and confirms the requirement for careful re-analysis of publicly available genomic data, with stringent quality control requirements. The study results highlight the emergence and dissemination of a distinct C2/*H*30Rx sub-cluster because of an introduction into Wales circa 1999 (sub-clade C2/*H*30Rx; 95% HPD: 1996 to 2001), which has been named GB-WLS.C2/*H*30Rx. The limited genomic diversity (median pair-wise distance of 11 SNPs) amongst a distinct GB-WLS.C2/*H*30Rx sub-cluster from North Wales (which emerged in 2009) suggests that the actual reservoir of infection reservoir was not confined to a single nosocomial setting. Analyses into local clusters and transmissions can be highly discriminatory and could become a routine part of surveillance programs with the generation of highly accurate short-reads, and high throughput, from Illumina’s short-read sequencing technologies.

Of particular concern is the high rates of ESBL carriage (66.7%) in ST131 bacteraemia isolates in Wales (clade C/*H*30), particularly those conferring resistance to 3GCs, from the *bla*_CTX-M-1_ (*n*=65/102, 63.7%). These rates are lower than studies in other jurisdictions; 95% of cephalosporin-resistant ST131 isolates in Australia, New Zealand and Singapore showed isolate carriage of *bla*_CTX-M-15_ or *bla*_CTX-M-27_(6) and 82.2% of cefotaxime-resistant UPEC isolates from South-West England carried *bla*_CTX-M_ variants(62). However, these study methodologies differed by specifically selecting 3GC-resistant isolates for inclusion, whereas this study is population-based and not biased by AMR selection. The rapid global emergence and sustained dominance of clade C/*H*30 ST131 and the characterisation of the unique ST131 Welsh sub-lineage (GB-WLS.C2/*H*30Rx) highlights the requirement for timely and continuous yearly genomic surveillance, which could facilitate rapid and targeted interventions, for a successful infection control, antimicrobial stewardship, and public health responses. The Office for National Statistics has previously collated mortality data where *E. coli* septicaemia or sepsis were explicitly mentioned on death certificates in Wales between 2001 and 2015(1). Taken together, this collated mortality data combined with genomic surveillance can support the National Health Service (NHS) in Wales by providing timely data for action on serious infections from both healthcare-and community-associated origins with little delay.

This study estimates the emergence of fluoroquinolone-resistant C/*H*30 ST131 circa 1995 (95% HPD: 1991 to 1998). This differs from the previously reported dates from Ben Zakour *et al*.(10), Stoesser *et al*.(61), and Kallonen *et al*.(4), that estimate 1987 (95% HPD: 1983 to 1992), 1982 (95% HPD: 1948 to 1995), and circa 1987, respectively. In contrast, after analysing 794 ST131 genomes, Ludden *et al*.(59) supports our findings and pinpoints the emergence of the fluoroquinolone-resistant C/*H*30 ancestor to 1992 (95 % HPD 1989 to 1994). While the posterior mean/median node heights for the clades vary between studies, it is important to recognise that the 95% HPD intervals overlap. The variation in study results is likely due to the enhanced methodology utilised, including: stringent quality control metrics, improved versions of tools and methods, use of a high quality clade C reference genome, the exclusion of clade B ST131 strains, and the inclusion of a fluoroquinolone sensitive clade C/*H*30 outgroup strain (CD306).

These highly discriminatory analyses reveal multiple introductions of sub-clade C2/*H*30Rx into Wales before an emergence circa 1999 (95% HPD: 1996 to 2001) of the unique clonal sub-lineage (GB-WLS.C2/*H*30Rx), which shares a CA of North American origin. These unique strains (GB-WLS.C2/*H*30Rx) were related with a median pair-wise SNP distance of 48 non-recombinogenic SNPs, which could indicate localised transmission with an unidentified infection reservoir. The CA to this unique strain (GB-WLS.C2/*H*30Rx) is distinguishable from that shared with the basal S125EC strain by an impairment of ferric enterobactin synthesis and transport due to a premature termination because of a Q130stop codon in *fepE*, and the loss of a region encompassing the capsular biosynthesis genes for a K5 capsular antigen. Notably, both enterobactin and the capsule are known UPEC virulence factors. Whether the inactivation of these genes resulted in a decrease in virulence remains to be elucidated and represents a research question for future investigation. For strains within GB-WLS.C2/*H*30Rx, region II of the capsular loci was replaced with a chloramphenicol acetyltransferase (CAT) and HAD-IA family hydrolase. CATs inactivate chloramphenicol by generating derivatives like 1-acetoxy chloramphenicol, 3-acetoxy chloramphenicol, or 1,3-diacetoxy chloramphenicol. These derivatives are unable to inhibit bacterial growth and survival as interruption of the ribosomal peptidyl-transferase is no longer possible(63, 64). Further, the identification of a local cluster within North Wales, MRCA emerged in 2009, with very closely related strains differing by a median of 11 non-recombinant pair-wise SNPs, suggests that there was possible direct transmission between these individuals. Although, the anonymisation of patient data limits confirmation of an actual nosocomial infection reservoir and evidence of negative (or positive) culture on admission would be required for any certainty.

The study design describes cases of bacteraemia, with confirmed blood cultures, caused by ST131 in Wales. This is likely a consequence of UTI treatment failure due to AMR, although further research is needed to establish links between confirmed blood and urine isolates. Therefore, this study may not represent the whole population structure of GB-WLS.C2/*H*30Rx in UTIs in Wales. In this circumstance, one would expect increased rates of AMR, and thus it is likely that the population structure of all GB-WLS.C2/*H*30Rx is to be less resistant than might be expected based upon the results reported here. Globally there is a necessity to acquire a deeper understanding of the population structure of UPEC, so that UPEC strains that are more likely to result in treatment failure and progress to bacteraemia can be identified as a risk factor. This can be achieved through the identification and tracking of genomic sequences (e.g. AMR determinants and virulence factors) as indicators for predicting phenotypic characteristics. One of the key strengths of our study was our ability to avoid a temporal or geographical bias in our dataset by contextualising the ST131 Welsh strains with global isolates. This lack of bias was reflected by our population expansion timeline which coincides with the initial detection of ST131 in the UK in 2003(4), before becoming the predominant clone (*n=*52/88, 59.1%) in the Northwest of England between 2004 and 2006(7). Previous studies of whole-genome SNP discovery for phylogenetics(6, 9, 10) have used the Bowtie 2(37, 65) or the SHRiMP(66) read aligner with FreeBayes(67) within the Nesoni pipeline (https://github.com/Victorian-Bioinformatics-Consortium/nesoni). However, SPANDx was utilised for the methodology employed in this study as it has been peer reviewed(46) and is under regular development, with v4.0.1 released on 02 April 2020. Additionally, previous quality assurance analyses using SPANDx have been reported(68) to ensure; a single mixed strain does not affect tree topology and phylogenetic inference, and the importance of assessing datasets for the presence of mixed strains prior to phylogenetic analyses.

### 8.1 Conclusion

Genomic epidemiological analyses on 142 ST131 strains associated with bacteraemia across Wales between 2013 and 2014 were performed using whole-genome sequencing. This research demonstrates the requirement to reanalyse publicly available genomic data, with stringent quality control, to improve the evolutionary trajectory of the ST131 clade C/*H*30 previously characterised. This study showed geographical clustering of sub-clade C2/*H*30Rx in North Wales; characterised by genotypic resistance to third-generation cephalosporins, fluoroquinolones, chloramphenicol, and nitrofurantoin. This emergence follows the introduction of a single sub-lineage into Wales circa 1999 and its expansion and persistence, which the authors have named GB-WLS.C2/*H*30Rx. This study highlights the need to incorporate whole-genome sequencing with epidemiological data and a ‘One Health’ approach to identify potential infection reservoirs in the environment, which will allow for the identification of ST131 transmission dynamics between healthcare settings and the community. This study displays a novel localised cluster of ST131 bacteraemia in Wales captured between 2013 and 2014. By gaining a detailed understanding of significant *E. coli* bacteraemia strains, it should be possible to develop targeted public health measures to reduce the risk of *E. coli* bacteraemia and act to combat the rise of antimicrobial resistance.

## Data Availability

The study sequences are available in the National Center for Biotechnology Information (NCBI) under BioProject accession number PRJNA729115. Raw Illumina sequence read data have been deposited to the NCBI sequence read archive [SRA (https://www.ncbi.nlm.nih.gov/sra)] under the accession numbers SRR14519411 to SRR14519567. A complete list of SRA accession numbers is available in Table S1 (available in the online version of this article). The high-quality draft assemblies have been deposited to GenBank under the accession numbers JAHBGJ000000000 to JAHBMG000000000, and JAHBRR000000000 to JAHBRT000000000. The programs used to analyse raw sequence reads for polymorphism discovery and whole-genome sequencing based phylogenetic reconstruction are available as described in the materials and methods. The authors confirm all supporting data, code, and protocols have been provided within the article or through supplementary data files.

## 9. Data Bibliography

1. White R.T. *et al*. BioProject PRJNA729115 (2021).
2. Kidsley A. K. *et al*. BioProject PRJNA627752 (2020).
3. Johnson, T. J. *et al*. BioProject PRJNA307507 (2016).
4. Johnson, T. J. *et al*. BioProject PRJNA311313 (2016).
5. Petty, N. K. *et al*. BioProject PRJEB2968 (2014).
6. Price, L. B. *et al*. BioProject PRJNA211153 (2013).
7. Andersen, P. S. *et al*. BioProject PRJNA218163 (2013).
8. Totsika, M. *et al*. BioProject PRJEA61443 (2011).
9. Toh H. *et al*., BioProject PRJDA19053 (2010).

## 10. Author statements

### 10.1 Authors and contributors

Conceptualisation: R.T.W. Investigation: R.T.W. Funding was acquired by T.R.C. and computational resources were supported by S.A.B. Formal analysis: R.T.W. Wet-lab experiments: M.J.B. and C.R.B. Data analysis: R.T.W. Data curation: central Specialist Antimicrobial Chemotherapy Unit (SACU) at Public Health Wales, University Hospital Wales, T.R.C., and R.T.W. Illumina sequencing was done at the Oxford Genomics Centre and MicrobesNG at the University of Birmingham. Supervision: T.R.C., B.M.F., and S.A.B. Writing (Original Draft Preparation): R.T.W. Writing (Review and Editing): R.T.W., M.J.B., C.R.B., J.M.A., M.W., L.S.J., R.A.H., M.M., M.M.A., B.M.F., T.R.C., and S.A.B. All authors have read and approved the final version of the manuscript.

### 10.4 Ethical approval

This work was undertaken on stored bacterial cultures and no additional clinical samples were collected from any persons to facilitate this study. Patient anonymity was maintained by pseudonymised data that went outside Public Health Wales.

## 10.5 Acknowledgements

We thank the Public Health Wales laboratories and the Specialist Antimicrobial Chemotherapy Unit at University Hospital Wales for their contributions from the national public health surveillance of bacteraemia cases in Wales. We acknowledge the facilities, and the scientific and technical assistance of staff at the Oxford Genomics Centre and MicrobesNG at the University of Birmingham. This research was supported by QRIScloud and by use of the Nectar Research Cloud. The Nectar Research Cloud is a collaborative Australian research platform supported by the National Collaborative Research Infrastructure Strategy (NCRIS). Sequence data are uploaded and stored on the centralised Cloud Infrastructure for Microbial Bioinformatics (MRC-CLIMB) server, which is funded by the Medical Research Council (MRC) (grant codes MR/L015080/1 and MR/T030062/1). The author would like to thank Derek Sarovich and Erin Price (GeneCology Research Centre at the University of the Sunshine Coast, and the Sunshine Coast Health Institute) and Thomas Cuddihy (QFAB Bioinformatics and Research Computing Centre, The University of Queensland) for high-performance computing support and helpful discussions about software functionality.

## Author notes

All supporting data, code and protocols have been provided within the article or through supplementary data files. Supplementary methods and supplementary tables are available with the online version of this article.

## 1.6 Abbreviations

3GCs: third-generation cephalosporins
AMR: antimicrobial resistance
BWA: Burrows– Wheeler Aligner
CA: common ancestor
*catB*: chloramphenicol-related O-acetyltransferase
CDS: coding sequence
CI: confidence interval
contigs: contiguous sequences
ECB: *Escherichia coli* bacteraemia
ESBLs: extended-spectrum *β*-lactamases
*febE*: ferric enterobactin transport protein
*fryC*: fructose-like permease IIC component
*fumC*: fumarate hydratase class II
GATK: Genome Analysis Tool Kit
HPD: highest posterior density
INDELS: insertions and deletions
IQR: interquartile range
IS: insertion sequences
MCC: maximum clade credibility
MCMC: Markov chain Monte Carlo
*mdh*: malate dehydrogenase
ML: maximum likelihood
MLST: multilocus sequence typing
NCBI: National Center for Biotechnology Information
NHS: National Health Service
NICE: National Institute for Clinical Excellence
PHW: Public Health Wales
RefSeq: Reference Sequence
SNPs: single-nucleotide polymorphisms
SACU: Specialist Antimicrobial Chemotherapy Unit
SRA: Sequence Read Archive
ST: sequence type
syn: synonymous
UK: United Kingdom
UPEC: Uropathogenic *Escherichia coli*
USA: United States of America
UTIs: urinary tract infections
WGS: whole-genome sequencing
XAT: xenobiotic acyltransferase

