## Supplemental Methods and Figures for "Genomic epidemiology reveals geographical clustering of multidrug-resistant *Escherichia coli* sequence type (ST)131 associated with bacteraemia in Wales, United Kingdom"

#### 1.1 Author names

Rhys T. White[1,2] https://orcid.org/0000-0001-6620-758X, Matthew J. Bull[3,4] https://orcid.org/0000-0002-8701-0417, Clare R. Barker[3] http://orcid.org/0000-0002-3276-5628, Julie M. Arnott[5,] Mandy Wootton[4] https://orcid.org/0000-0002-2227-3355, Lim S. Jones[4], Robin A. Howe[4], Mari Morgan[5], Melinda M. Ashcroft[6] https://orcid.org/0000-0001-9157-4533, Brian M. Forde[7] https://orcid.org/0000-0002-2264-4785, Thomas R. Connor[3*] https://orcid.org/0000-0003-2394-6504, Scott A. Beatson[1,2*] https://orcid.org/0000-0002-1806-3283

#### 1.2 Affiliation

[1]School of Chemistry and Molecular Biosciences and Australian Infectious Disease Research Centre, The University of Queensland, Brisbane, Queensland 4072, Australia

[2]Australian Centre for Ecogenomics, The University of Queensland, Brisbane, Queensland 4072, Australia

[3]Microbiomes, Microbes and Informatics Group, Organisms and Environment Division, School of Biosciences, Cardiff University, Cardiff, Wales CF10 3AX, United Kingdom

[4]Public Health Wales Microbiology, University Hospital Wales, Cardiff, Wales CF14 4XW, United Kingdom

[5]Healthcare Associated Infection, Antimicrobial Resistance & Prescribing Programme (HARP), Public Health Wales, 2 Capital Quarter, Tyndall Street, Cardiff, Wales CF10 4BZ, United Kingdom

[6]Department of Microbiology and Immunology, The University of Melbourne at The Peter Doherty Institute for Infection and Immunity, Melbourne, Victoria, Australia

[7]The University of Queensland, UQ Centre for Clinical Research (UQCCR) and Australian Infectious Disease Research Centre, Royal Brisbane & Women's Hospital Campus, Herston, Queensland 4029, Australia

#### 1.3 Corresponding authors

Thomas R. Connor;


Scott A. Beatson;


**<u>This file includes:</u>**

Supplementary Methods: Sections 2.1 through to 2.4

Supplementary Figure S1. Deaths registered in each calendar year in Wales involving *Escherichia coli* septicaemia between 2001 and 2015.

Supplementary Figure S2. Population estimates by lower layer super output areas, 2014.

Supplementary Figure S3. Nucleotide comparisons between key sequence type determining housekeeping genes within the reference chromosome EC958.

Supplementary Figure S4. Maximum likelihood phylogenetic analysis representing global *Escherichia coli* sequence type (ST)131.

Supplementary Figure S5. Maximum parsimony phylogeny of clade C/*H*30 *Escherichia coli* sequence type (ST)131 isolates plotted against *β*-lactam resistance complement.

Supplementary Figure S6. Evolutionary reconstruction of clade C/*H*30 *Escherichia coli* sequence type (ST)131.

### 2. Supplementary Methods

#### 2.1 Quality control of sequence data for the 157 Welsh strains

The FastQC package v0.11.8 (http://www.bioinformatics.babraham.ac.uk/projects/fastqc/) was used to generate quality statistics for the paired-end reads, which were aggregated into a single report and visualised using MultiQC v1.7(1). Kraken v2.0.7-beta(2) was then used to screen the raw Illumina sequencing data for contamination against the National Center for Biotechnology Information (NCBI) RefSeq database(3). Raw reads were filtered using Trimmomatic v0.36(4) by removing low-quality bases and read pairs together with Illumina adaptor sequences (settings: LEADING:10 TRAILING:10 MINLEN:50 HEADCROP:10). The average sequence coverage depth was estimated using the Burrows–Wheeler Aligner v0.7.15(5); SAMtools v1.2(6); Picard v2.7.1 (https://github.com/broadinstitute/picard); the Genome Analysis Tool Kit v3.2-2 (GATK)(7, 8); BEDTools v2.18.2(9); and SNPEff v4.1(10) as implemented in SPANDx v3.2(11). In brief, the trimmed reads were mapped to the complete chromosome of *E. coli* ST131 strain EC958 (GenBank: HG941718); which was isolated from the urine of an 8-year-old girl presenting in the community in March 2005 in the United Kingdom(12). We identified and excluded the sequence data for 15 isolates from further analysis based on the sequencing coverage below 20-fold (Table S3).

#### 2.2 *In silico* gene typing

MLST v2.19.0 (https://github.com/tseemann/mlst) with default settings was used to characterise the multi-locus sequence type (MLST) of the 142 strains by querying the high-quality draft assemblies against the *E. coli* MLST allelic profiles hosted on PubMLST(13, 14). ABRicate v0.9.7 (https://github.com/tseemann/abricate) was used to screen the high-quality draft assemblies for O and H-antigens, acquired antimicrobial resistance genes, and bacterial plasmid replicons using the EcOH(15), ARG-ANNOT(16), and PlasmidFinder(17) databases, respectively (last updated 07th September 2019). PointFinder(18) was used to screen high-quality draft assemblies for chromosomal point mutations, particularly in the QRDR of *gyrA*, *gyrB*, *parC*, and *parE* genes(19, 20). The K-antigen and *fimH* allele were characterised using Kaptive v0.4(21) (default settings) against a custom *E. coli* database comprising of known capsule antigens from complete genomes available on NCBI and FimTyper 1.0 (https://cge.cbs.dtu.dk/services/FimTyper/) with default parameters, respectively.

#### 2.3 Assembly based ST131 phylogeny

We initially aimed to place our Welsh isolates (*n*=142) within the context of ST131 isolates sampled globally (*n*=208). The generation of a core-genome (based on homology) multi-alignment, and identification of single-nucleotide polymorphisms (SNPs) was performed with Parsnp v1.2(22), utilising the PhiPack recombination filter(23). A total of 13,854 (13,758 non-recombinant) core-genome SNPs relative to the reference chromosome EC958 were identified from a 2,575,140 bp core-genome alignment. Finally, RAxML v8.2.10(24) with GTR-GAMMA correction generated a Maximum Likelihood (ML) phylogeny thorough optimisation of the 20 distinct randomized Maximum Parsimony trees generated from the 13,758 non-recombinant core-genome SNP alignment.

#### 2.4 Identifying and removing strain mixtures from the clade C/*H*30 ST131 phylogeny

A total of 225 strains representing our previously published collection [*n*=123, including 16 complete genomes (including EC958)] and our Welsh collection (*n*=102) were identified as either sub-clade C1/*H*30R or C2/*H*30Rx ST131 as described above. To assess the 225 strains for the presence of strain mixtures, paired-end reads were mapped onto the chromosome of EC958 using SPANDx to generate annotated SNPs and insertions and deletions (INDELs) matrices. Heterozygous SNPs in each genome were identified from GATK UnifiedGenotyper VCF output. A total of seven genomes were classified as strain mixtures based on the orthologous SNP alignment containing ~3% or more heterozygous SNPs sites and were subsequently removed from the dataset, leaving 218 genomes. These includes strains: JJ1908 (*n*=309/7,592, 4.1%); P146EC (*n*=299/7,592, 3.9%); S43EC (*n*=289/7,592, 3.8%); ZH164 (*n*=285/7,592, 3.8%); S39EC (*n*=278/7,592, 3.7%); G150 (*n*=272/7,592, 3.6%); and S30EC (*n*=210/7,592, 2.8%).

### 3. Supplementary Figures

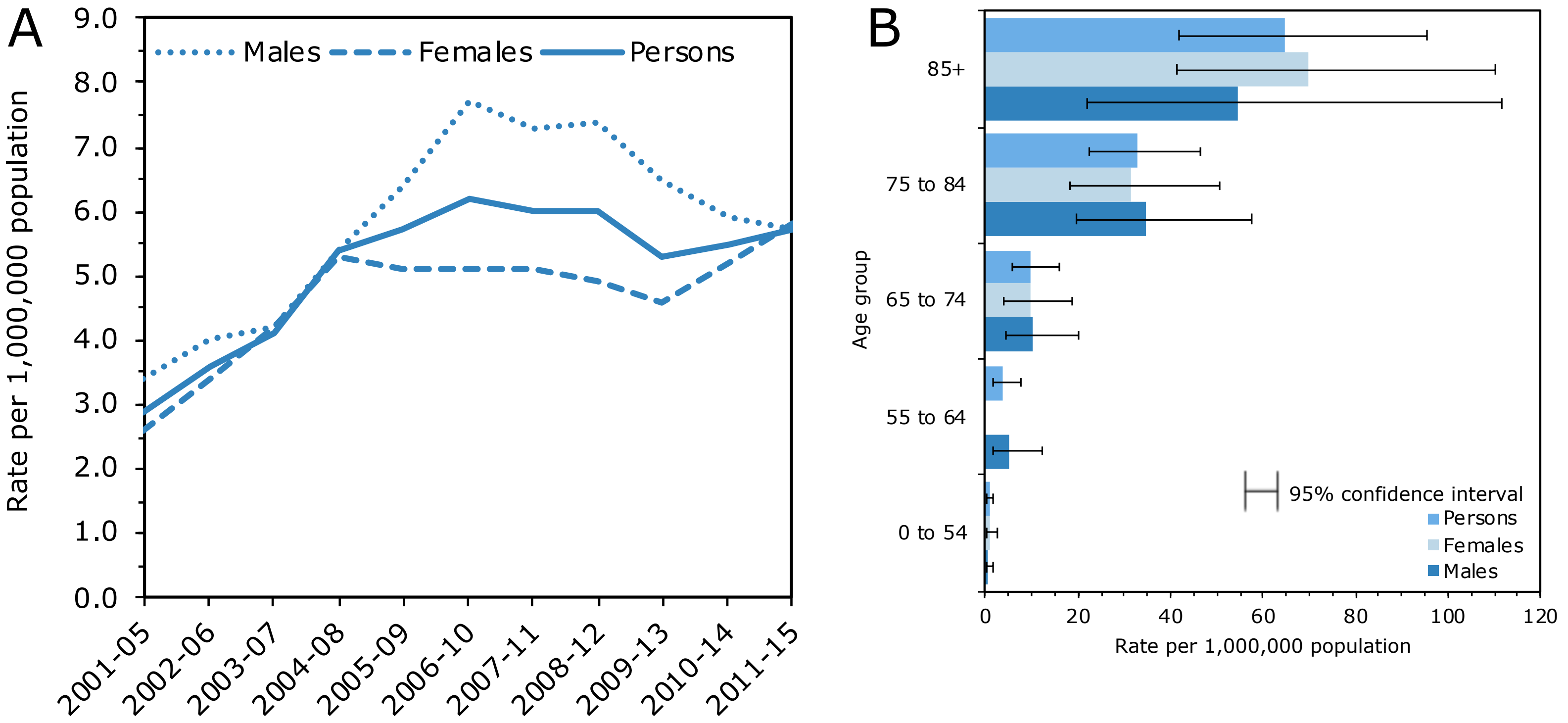


**Figure S1. Deaths registered in each calendar year in Wales involving *Escherichia coli* septicaemia between 2001 and 2015.** Due to small numbers of deaths for individual years, deaths were pooled into 5-year periods to calculate more robust rates. Figures are based on postcode boundaries as of May 2016 and exclude deaths of non-residents. Statistically significant differences between rates were assessed using 95% confidence intervals (CI). (A) Age-standardised mortality 5-year rolling rates per million population. 95% CI are not displayed as there is no significant difference between sexes. (B) Age-specific mortality 5-year rolling rates per million population. Adapted from "Deaths involving *E. coli* septicaemia, deaths registered in Wales between 2001 and 2015 [Online]" by the Office for National Statistics. Available at: https://www.ons.gov.uk/peoplepopulationandcommunity/birthsdeathsandmarriages/deaths/adhocs/006005deathsinvolvingecolisepticaemiadeathsregisteredinwalesbetween2001and2015 [Accessed 31st January 2017]. (2016). Copyright © 2016 by Office for National Statistics.

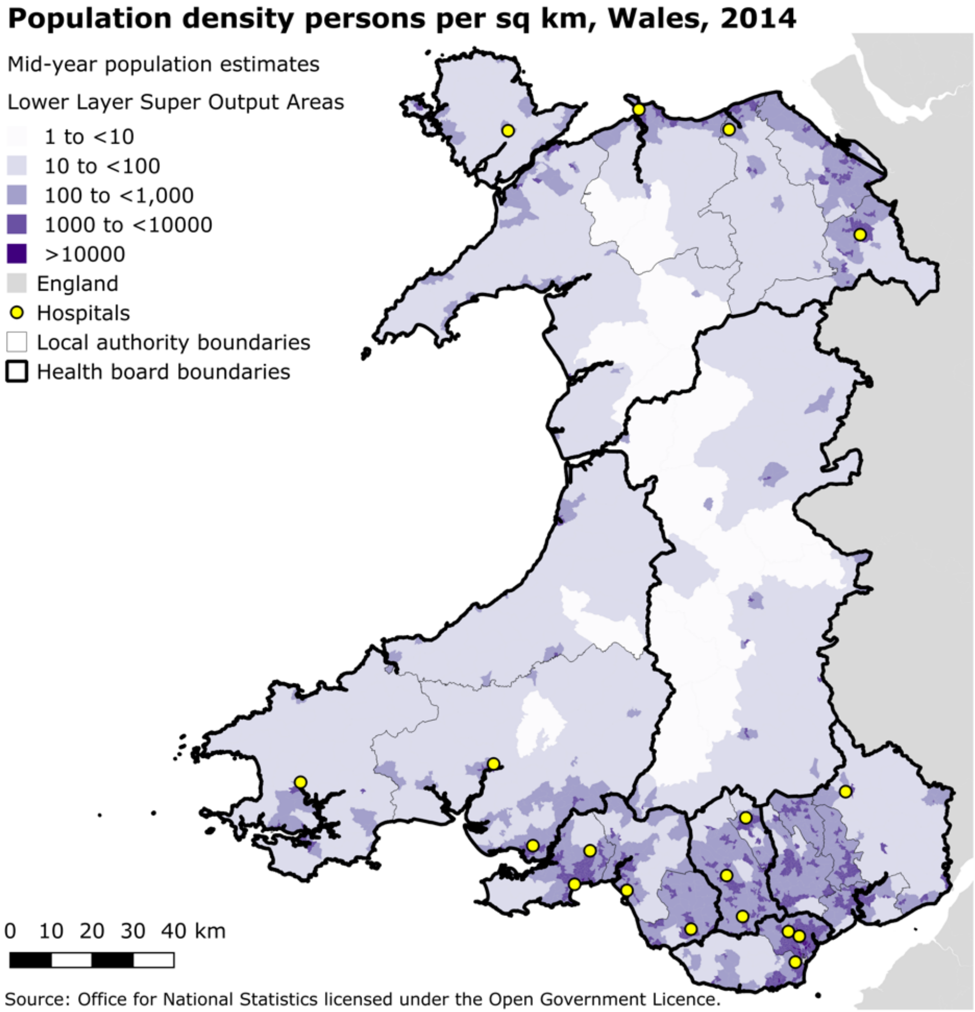


**Figure S2. Population estimates by lower layer super output areas, 2014.** The population Wales is expressed as population per square kilometre of land.

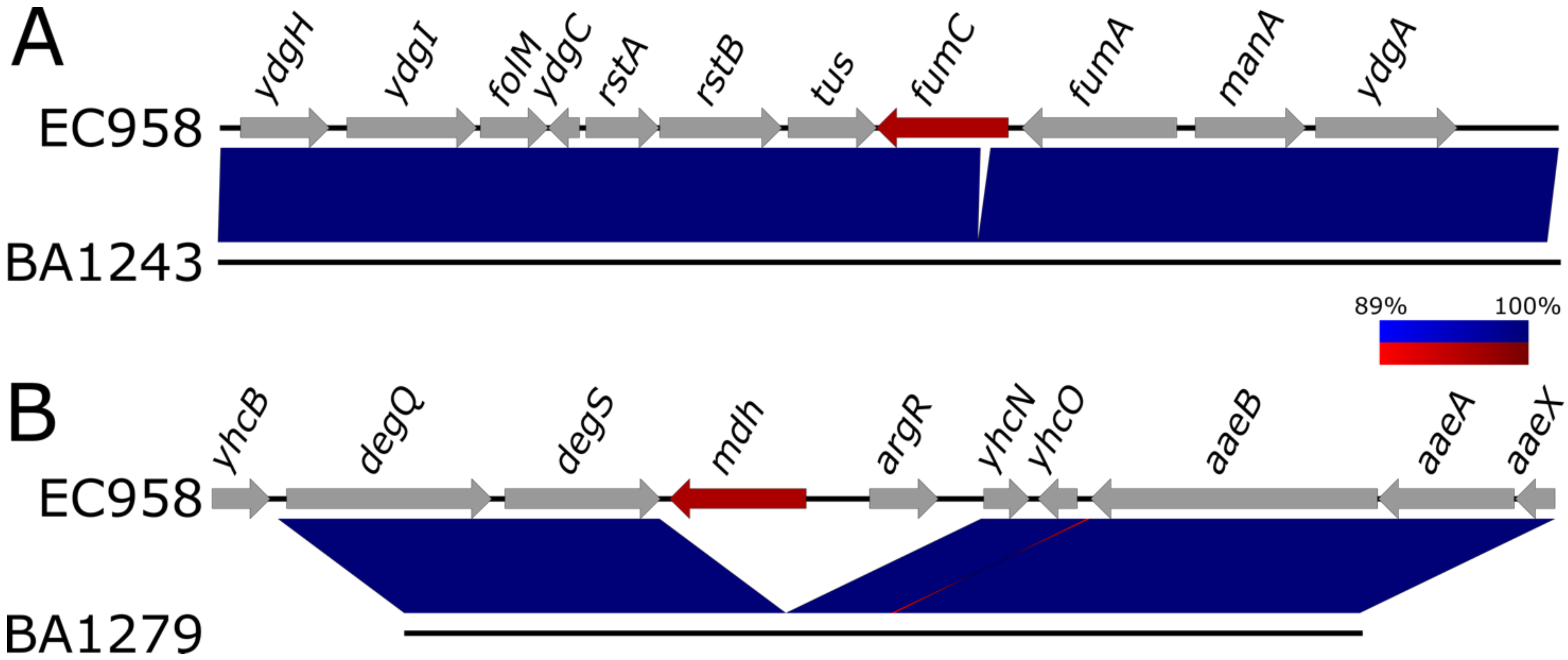


**Figure S3. Nucleotide comparisons between key sequence type determining housekeeping genes within the reference chromosome EC958**. Blue shading indicates nucleotide identity (red, inverted regions) between sequences according to BLASTn (89 to 100%). Key housekeeping genes are indicated in red, other CDSs in grey. Image created using EasyFig(25).

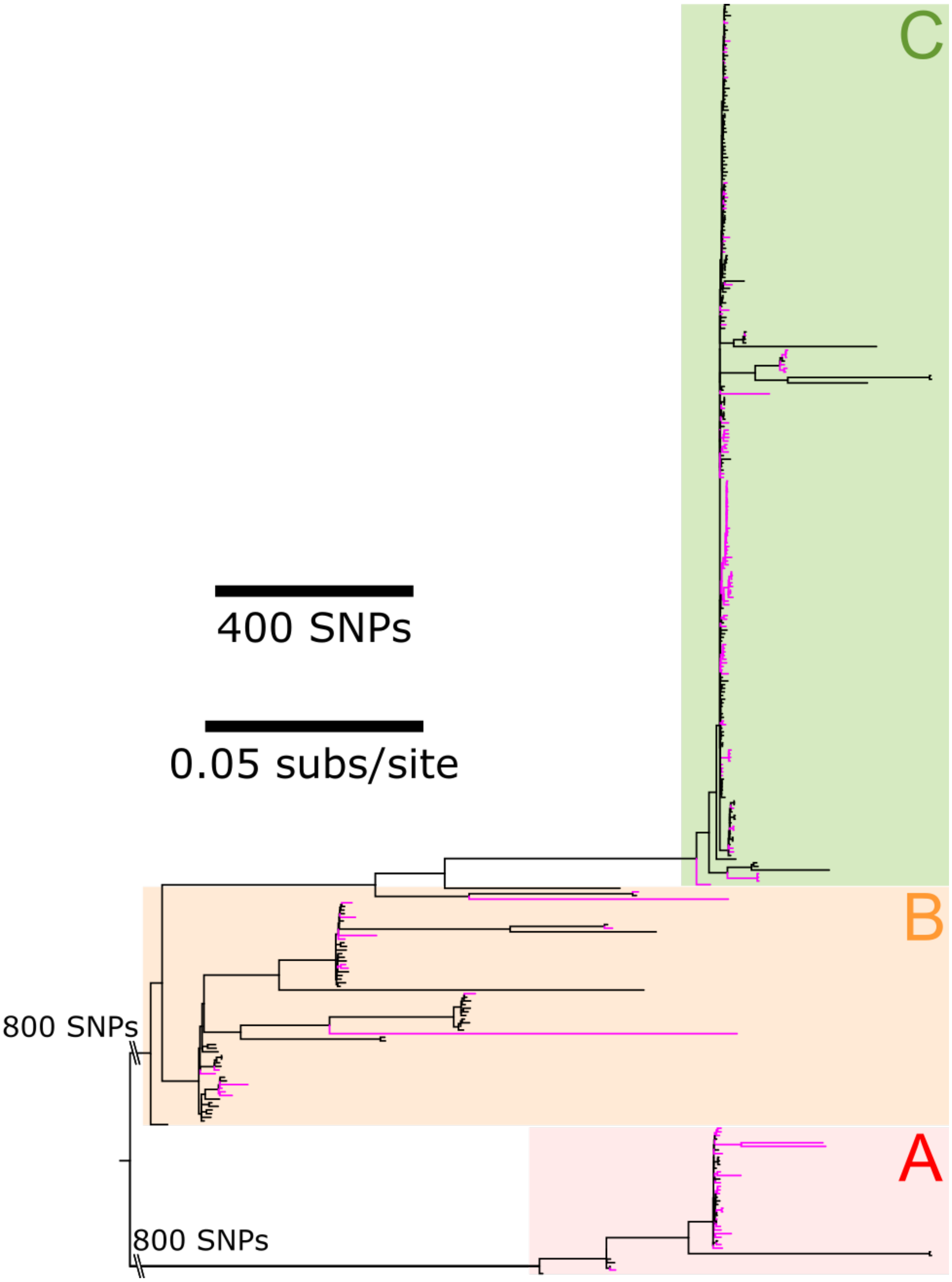


**Figure S4. Maximum likelihood phylogenetic analysis representing global *Escherichia coli* sequence type (ST)131.** Phylogeny is inferred from 13,758 non-recombinant core-genome single nucleotide polymorphisms (SNPs) relative to the reference chromosome EC958. SNPs were identified with Parsnp with the PhiPack recombination filter and represent a 2,575,140 bp core-genome. Phylogenetic tree is rooted according to the midpoint. Branch lengths represent SNP distances or nucleotide substitutions per site as indicated by the scale bars. The major ST131 phylogenetic clades are indicated: A = red, B = yellow, and C = green. Strains from Wales (*n*=142) are highlighted in magenta.

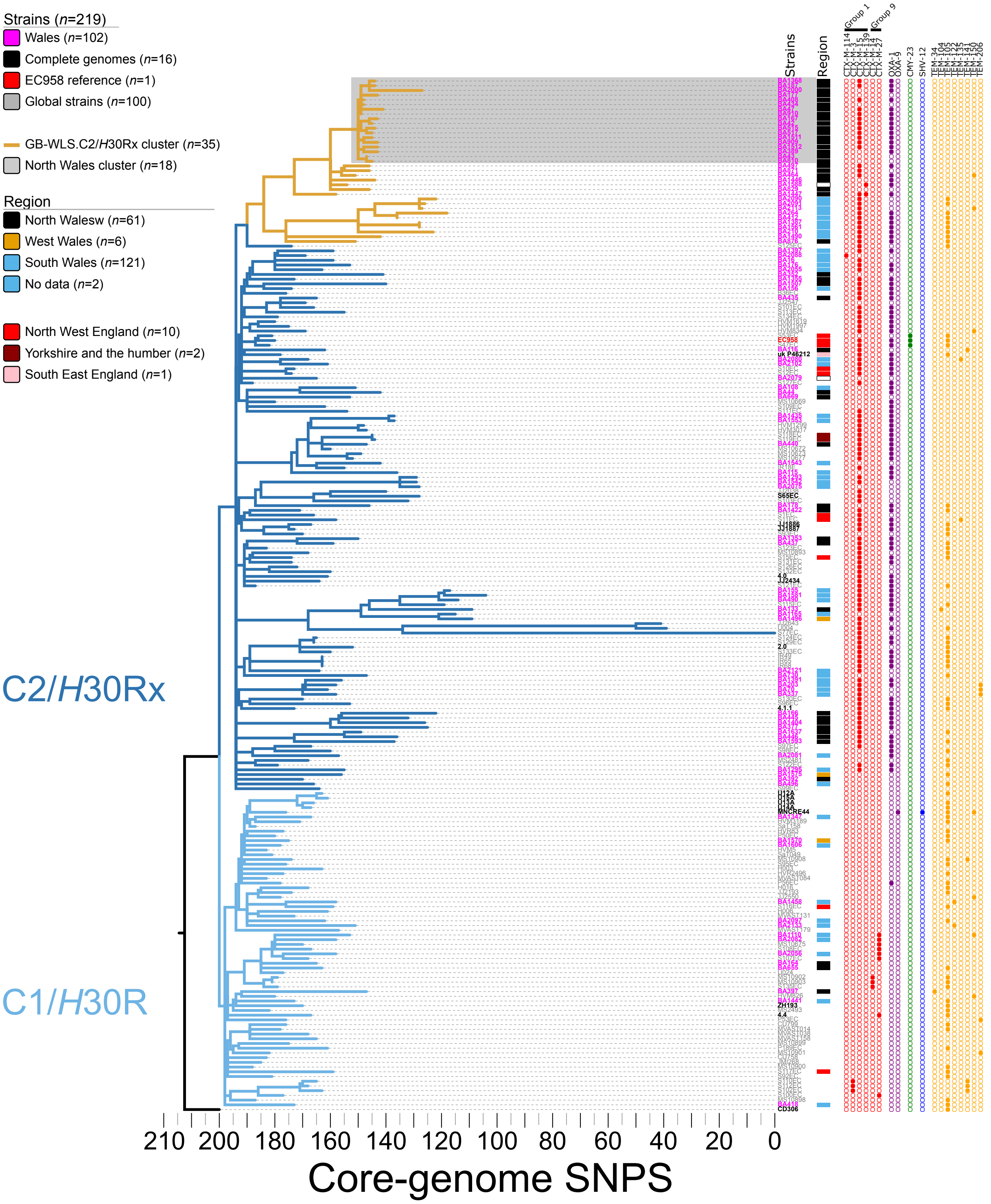


**Figure S5. Maximum parsimony phylogeny of clade C/*H*30 *Escherichia coli* sequence type (ST)131 isolates plotted against *β*-lactam resistance complement.** Phylogeny inferred from 4,354 non-recombinant orthologous biallelic core-genome single-nucleotide polymorphisms (SNPs) from 219 strains. Moderate recombination SNP density filtering in SPANDx (excluded regions with ≥3 SNPs in a 100 bp window). SNPs are derived from read mapping to the reference chromosome EC958 (GenBank HG941718). Phylogenetic trees are rooted according to the CD306 (GenBank: CP013831) outgroup. Branch lengths represent SNP distances indicated by the scale bar. The consistency index is 0.94.

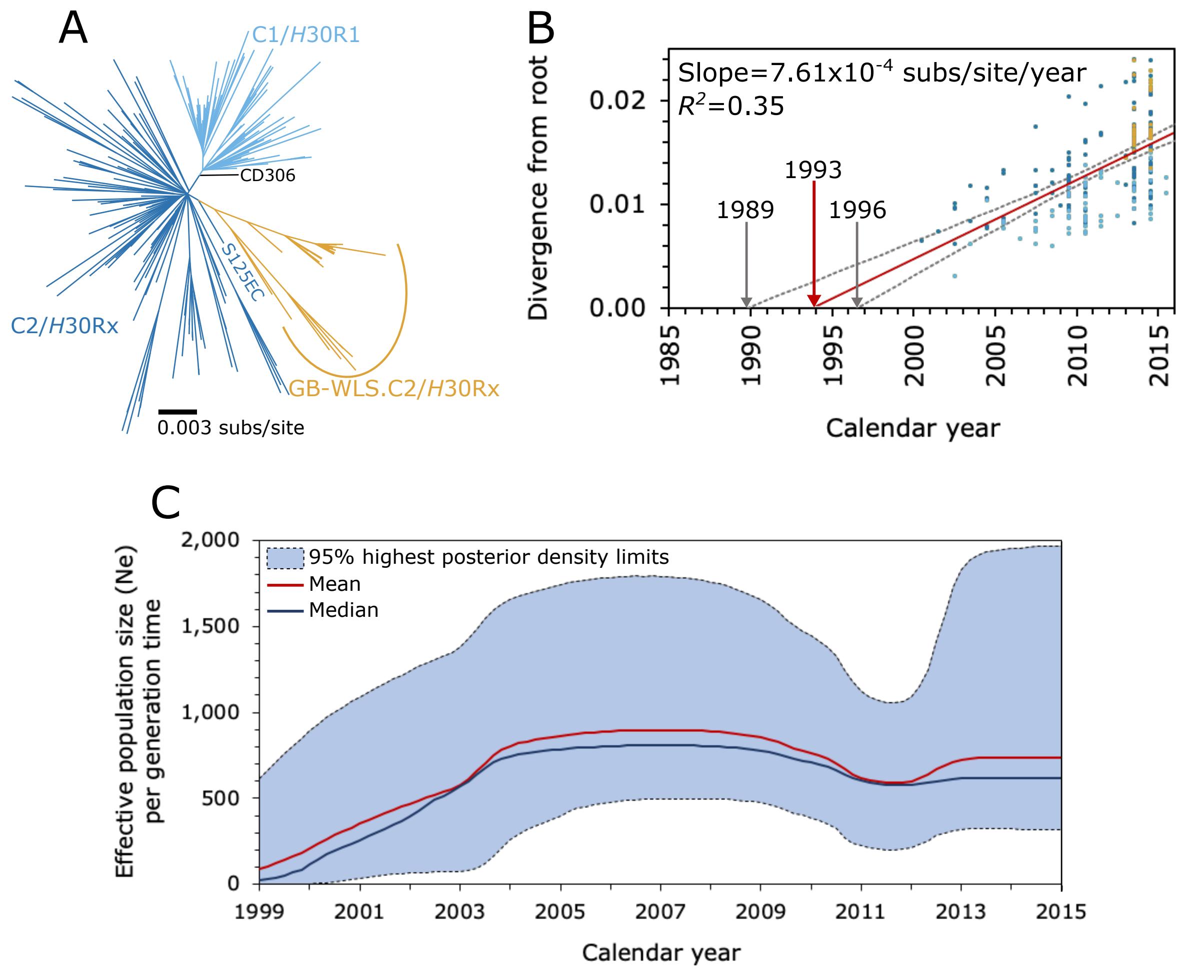


**Figure S6. Evolutionary reconstruction of clade C/*H*30 *Escherichia coli* sequence type (ST)131.** (A) Maximum likelihood phylogeny of 215 clade C/*H*30 isolates inferred from 4,150 non-recombinant orthologous biallelic core-genome single-nucleotide polymorphisms (SNPs). Moderate recombination SNP density filtering in SPANDx (excluded regions with ≥3 SNPs in a 100 bp window). SNPs are derived from read mapping to the reference chromosome EC958 (GenBank: HG941718). Branch lengths represent nucleotide substitutions per site as indicated by the scale bar. (B) Linear regression of root-to-tip genetic distance plotted against year of collection as implemented in TempEST. The substitution rate for the tree in A is indicated by the slope of the solid red regression line supported by 95% confidence intervals (grey dashed lines). Trees in A and B were rooted according to the *E. coli* CD306 (GenBank: CP013831) outgroup. (C) A Bayesian Skyline plot showing the predicted demographic changes of the ST131 clade C/*H*30 population since 1999.
